# *NXPE1* alters the sialoglycome by acetylating sialic acids in the human colon

**DOI:** 10.1101/2024.09.25.24314083

**Authors:** Bum Seok Lee, Ashley Cook, Surojit Sur, Laura Dobbyn, Maria Popoli, Sana Khalili, Shibin Zhou, Chetan Bettegowda, Nickolas Papadopoulos, Kathy Gabrielson, Phillip Buckhaults, Bert Vogelstein, Kenneth W Kinzler, Nicolas Wyhs

**Affiliations:** Ludwig Center for Cancer Genetics and Therapeutics, Johns Hopkins University School of Medicine, Baltimore, MD 21205, USA; Cellular and Molecular Medicine Graduate Program, Johns Hopkins University School of Medicine, Baltimore, MD 21205, USA; Department of Oncology, Johns Hopkins Medical Institutions, Baltimore, MD 21287, USA; Department of Molecular and Comparative Pathobiology, Johns Hopkins University School of Medicine, Baltimore, MD 21205, USA; College of Pharmacy, University of South Carolina, Columbia, SC 29208, USA; Department of Neurosurgery, Johns Hopkins University School of Medicine, Baltimore, MD 21205, USA; Howard Hughes Medical Institute, Johns Hopkins University School of Medicine, Baltimore, MD 21205, USA; Sol Goldman Pancreatic Cancer Research Center, Johns Hopkins University School of Medicine, Baltimore, MD 21205, USA

## Abstract

Mild periodic acid Schiff staining (mPAS) of human colonic tissue has been used to answer a variety of fundamental questions in germline and somatic genetics. mPAS staining is known to reflect sialic acid O-acetylation, but a full accounting of the genes contributing to sialoglycome diversity is incomplete. Using haplotypes derived from whole genome sequencing, we identified a region on chromosome 11 that is associated with inherited differences in mPAS staining. Of the genes in this region, only *NXPE1* haplotypes correlated perfectly with mPAS staining in the original cohort used for whole genome sequencing, as well as in a validation cohort. Transcriptomic analysis suggested that one common allele of *NXPE1*, with a single base pair substitution in its promoter region, greatly reduces expression of the gene. Genetic manipulation of *NXPE1* expression confirmed this conclusion and caused changes to modified sialic acid levels. Finally, high-performance liquid chromatography (HPLC) confirms that enzymatically active NXPE1 is capable of transferring an acetyl group from acetyl coenzyme A to sialic acid *in vitro*. These findings suggest that *NXPE1* is the long-sought gene responsible for differences in colon mPAS staining and may be the prototype of a new family of sialic acid O-acetylation-modifying genes.

## Introduction

Since 1946, pathologists have used periodic acid Schiff staining (PAS) to look for mucins - proteins that are heavily glycosylated and are a large constituent of mucus - in various human tissues^1^. Major constituents of the terminal end of glycosylated proteins, as well as glycosylated lipids, are sialic acids which, in turn, are post-translationally modified and together constitute the sialoglycome^2–4^. Such modifications are important as they can abrogate or encourage binding to other cellular constituents depending on the context of the interaction in question^3^. This impacts a number of cellular processes, including host-pathogen interactions and cell differentiation during development^5^. A variant of PAS staining, called mild periodic acid Schiff staining (mPAS), can distinguish between O-acetylated (mPAS negative) and non-O-acetylated (mPAS positive) sialic acids in tissues, including the colon^5–15^. Previous studies have demonstrated that mPAS staining patterns follow Hardy-Weinberg patterns in normal human colon tissue. Moreover, the number of sporadically positive crypts in otherwise negatively staining individuals increases with age, location within the colon, a history of radiation therapy, and chronic inflammatory states^11,16–20^. These somatically acquired changes in staining have been used to shed light on a number of fundamental questions, including the nature of stem cells and the rate of somatic mutation in normal colonic epithelium^21–25^. Uniform positive mPAS staining varies with racial background, with individuals of Asian descent staining positively more frequently than those of European or African descent^18^. Inbred murine models do not demonstrate variable staining, and are uniformly mPAS positive^26,27^.

These observations support the idea that an unknown autosomal dominant human gene is responsible for the differential O-acetylation that underlies mPAS staining^17,18^. To date, the only known human sialic acid O-acetyltransferase (SOAT) is encoded by the gene *CASD1*, but its correlation with human colon mPAS staining has not been investigated^28,29^. Given that acetylation modification occurs at many sites on sialic acid in different tissues, several other genes that control O-acetylation likely remain to be discovered^3,30,31^. We here describe studies suggesting that one such gene is neurexophilin and PC-esterase domain family member 1 (*NXPE1*).

## Results

### Whole genome sequencing identifies a haplotype on chromosome 11 associated with mPAS staining

Whole genome sequencing was performed on DNA from the colonic tissue of 21 normal individuals: 8 were negative by mPAS staining, 8 were positive, and another 5 were negative with at least one positive crypt among thousands of non-staining crypts. Such patients were presumed to be of heterozygous genotype for the unknown gene of interest (Figure 1A). The purpose of sequencing was to identify single-nucleotide polymorphisms (SNPs) or small insertions or deletions (indels) that correlated with mPAS staining (Table S1). mPAS staining is primarily localized to goblet cells within normal colorectal crypts (Figure 1A). An initial genome-wide review identified 202 SNPs with two or fewer genotype/phenotype mismatches in the 21 patients examined, with chromosome 11 harboring 90 such SNPs, far more than any other chromosome (Figures 1B and S1, Table S2). A more stringent analysis revealed that all 17 SNPs that showed perfect concordance with the mPAS phenotype were on chromosome 11 and enriched in a small region on chromosome 11q23.2 (chr11:114,298,921 to chr11:114,446,104) (orange bars in Figure 1B, Green dots in Figure S1). An expanded illustration of this 173kb region is shown in Figures 1C and S2, and shows a strong linkage disequilibrium (r^2^ values > 0.8) around the promoter region of the gene *NXPE1*.

**Figure 1:**
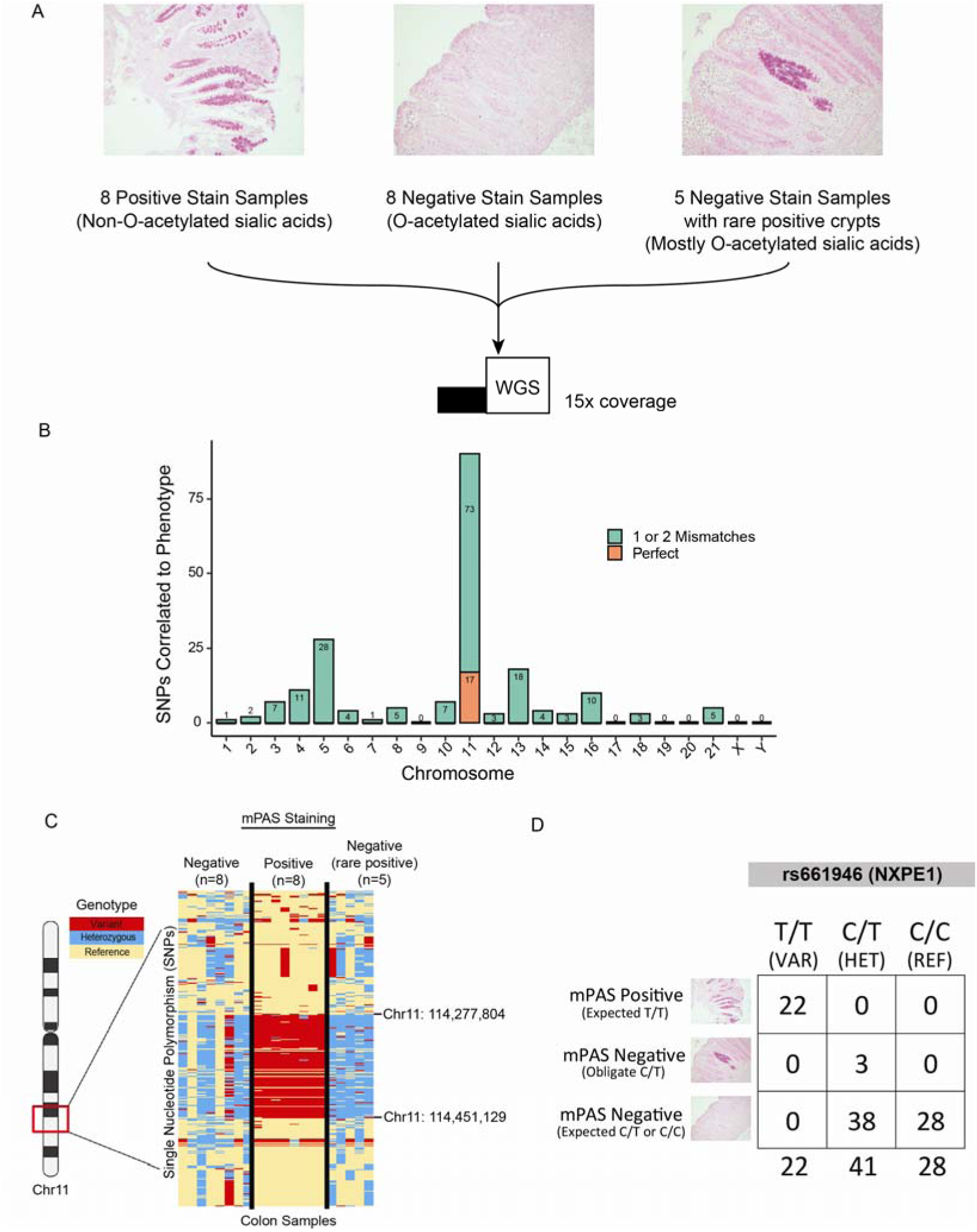
WGS identifies haplotype on chromosome 11 associated with colorectal sialic acid acetylation status based on mPAS staining. **A)** Schematic showing samples and NGS strategy employed to identify regions associated with colon mPAS staining. Negative staining samples with a rare positive crypt were presumed to be of heterozygous genotype, where a spontaneous loss of heterozygosity in a stem cell caused positive staining of a crypt (right). **B)** Bar plot showing the number of SNPs correlated with the mPAS phenotype by chromosome as evaluated by WGS. Only SNPs on chromosome 11 contained perfect matches between genotype and phenotype in all samples. **C)** Region on chromosome 11q23.2 identified by whole genome sequencing as highly associated with mPAS staining. Samples that largely stain negative but contain rare positive crypts are suspected of having a heterozygous genotype and are listed separately (right side). SNPs are shown on the y-axis based on their location on chromosome 11. Formal linkage disequilibrium analysis of this region is shown in figure S2. **D)** Independent validation of WGS results. 3×3 table showing genotype for SNP rs661946 (located in the promoter of *NXPE1*) and mPAS staining phenotype on a set of 91 normal colon tissue samples. Allele frequencies are consistent with Hardy-Weinberg equilibrium (□□□Haldane Exact = 0.45). Fisher’s Exact Test for the genotype-phenotype relationship yields a p < 2.2e-16.

### Targeted sequencing implicates *NXPE1* as the gene most tightly associated with colorectal sialic acid mPAS staining status

Four protein coding genes reside in the region described above: *NXPE1*, neurexophilin and PC-esterase domain family member 4 (*NXPE4*), RNA binding motif protein 7 (*RBM7*) and RNA exonuclease 2 (*REXO2*). A review of the 17 perfectly correlated SNPs showed none were in coding regions, four were upstream of either *REXO2* or *NXPE1*, and the others were in intergenic or intronic regions, or downstream, of one of the four genes (Figure S2). To extend these results, we obtained 91 new samples of normal colonic tissue and stained them to assess mPAS status. Targeted sequencing of the four SNPs upstream of *REXO2* and *NXPE1* showed that only one of them (rs661946) maintained a perfect match (91/91) between the observed phenotypic staining by mPAS and the genotype (Figure 1D and Table S3). This SNP was only 6 bp upstream of the transcriptional start site of NXPE1, presumably part of its promoter, and exists as part of or immediately adjacent to at least three transcription factor binding motifs (ETS-2, SOX1 and GR-α)^32–34^. The observed frequencies for this biallelic SNP (C and T at 53% and 47%, respectively) are consistent with Hardy-Weinberg equilibrium (Haldane’s Exact Test). They are also consistent with the published minor allele frequency (MAF) of mPAS positive staining for individuals of East Asian descent, the location from which most of the 91 samples originated^18,35^. The only other SNP significantly associated with mPAS staining was rs561722 (*NXPE2P1*), but it still contained 3/91 mismatches (Table S3).

### Structural similarities of *NXPE1* to genes known to O-acetylate sialic acid

*NXPE1* and *NXPE4* have been reported as containing secondary structural similarities to known sialic acid O-acetyltransferases from other species^36^. *NXPE1* contains two motifs, Gly-Asp-Ser (GDS) and Asp-X-X-His (DXXH), with identical amino acid sequence to the catalytic site of the only known human SOAT, the protein CASD1 (expect value=0.004, 23.9% identity)(Figures 2A and 2B). Like CASD1, NXPE1 protein lacks the canonical glycine and asparagine in the GDSL/SGNH fold present in SOATs from other species, making it part of the GDSL/SGNH-like acyl-esterase family^29,36^. That said, the catalytic sites of NXPE1 do contain similarities to the viral homologs of SOAT proteins found in influenza C/JHG/66 virus, Isavirus salaris, bovine torovirus strain Breda 2, and human coronavirus OC43 (Figures 2A and 2B bottom)^29,37,38^. These sequence similarities are also conserved among other members of the *neurexophilin/NXPE* family including *NXPE4*, though they display a different, His-Pro-Pro, sequence for their histidine containing motif (Figure 2B)^36,39^. *In silico* 3D structure prediction of NXPE1 co-locates Asp526 and His529 adjacent to the Ser355, creating a hydrogen bond between the histidine and serine residues similar to the catalytically active serine (Ser94) in CASD1 (Figures 2C and 2D). NXPE1 Ser355 is predicted to be exposed to solvent and to reside in the intra-endoplasmic reticulum/Golgi space (where glycan addition and modification occurs), suggesting these three residues (Asp526, His529, and Ser355) are likely important for catalytic activity of the protein (Figures 2D and S3)^40,41^.

**Figure 2:**
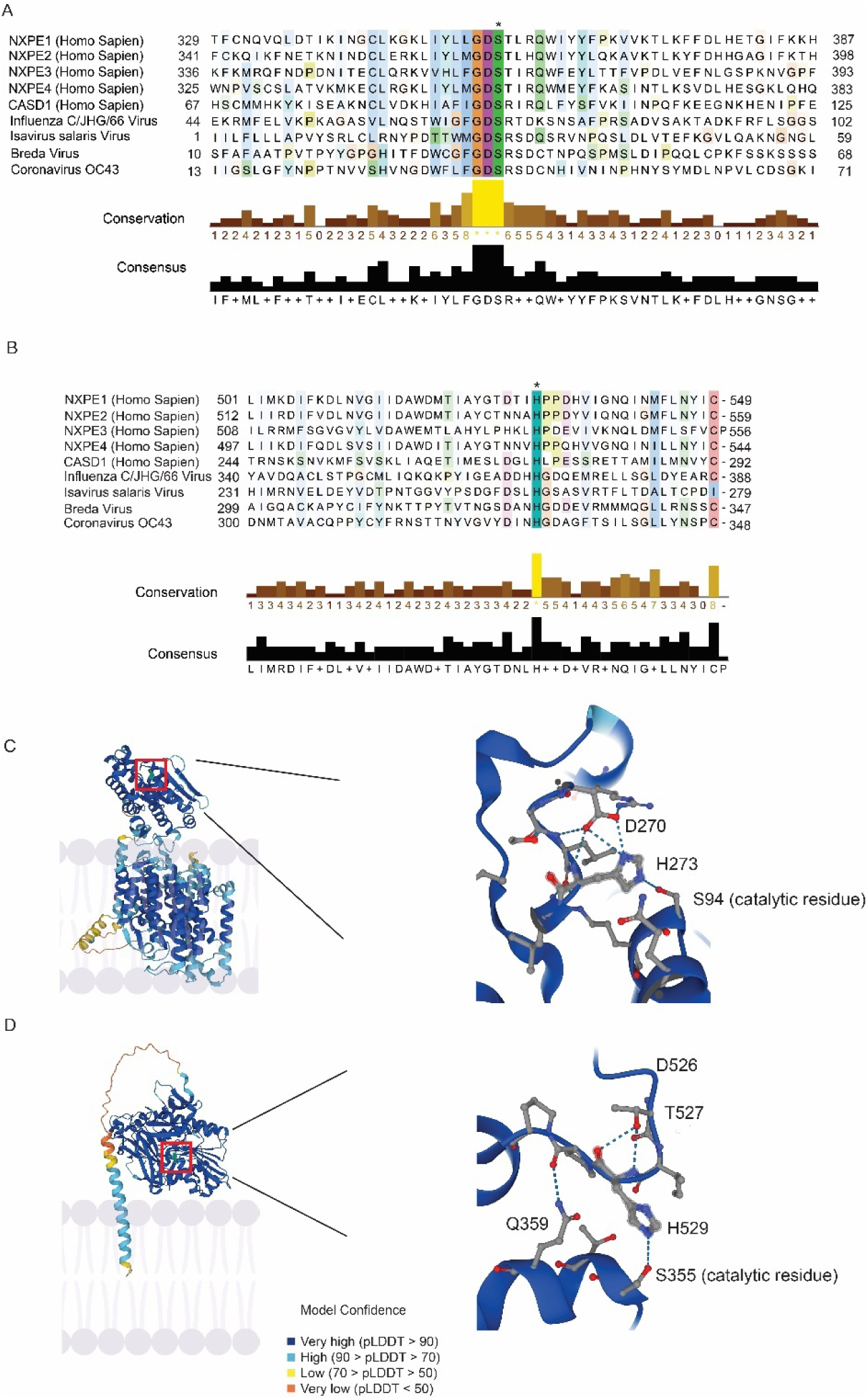
NXPE1 protein sequence and 3D structure have close homology to known sialic acid-O-acetyltransferases. *NXPE1* and *NXPE4* are part of a small family of proteins known as neurexophilin/NXPE, with 4 known members. The *NXPE1* gene codes for a 547 amino acid protein predicted to contain a N-terminal transmembrane domain, with the majority of the protein residing on the extracellular portion of the cell. **A)** Multiple sequence alignment of amino acid sequences near the serine and **B)** aspartate-histidine catalytic sites of NXPE family members with known sialic acid-O-acetyltransferases. NXPE1 (Q8N323.2) was compared to human proteins neurexophilin and PC-esterase domain family member 2 (NXPE2) (Q96DL1.2), neurexophilin and PC-esterase domain family member 3 (NXPE3) (Q969Y0.1), NXPE4 (Q6UWF7.1) and CASD1 (Q96PB1.1), as well as viral SOAT proteins from influenza C virus (C/Johannesburg/1/66) (P07975.1), Isavirus salaris (AAL34465.1), Breda virus (CAA71819.1), Human coronavirus OC43 (P30215.1). The active site residues are indicated with an *. Conservation shown above is a quantification of the alignment of residues as determined by pre-defined physio-chemical properties. Consensus refers to the percentage of which the most common residue was conserved. **C)** *In silico* predicted structure of CASD1 and **D)** NXPE1 showing co-localization of catalytic site serine from the Gly-Asp-Ser (GDS) motif with catalytic site histidine from the DXXH motif. All structures shown in this figure are predictions based on AlphaFold accessed 11/2023.

### *NXPE1* protein levels correlate with sialic acid mPAS status and to the staining of sialic acid binding lectins

We developed immunohistochemical (IHC) assays for NXPE1, NXPE4, and CASD1 (the only known human SOAT) and evaluated their expression in colonic tissue. All three proteins showed a cytoplasmic granular immunostaining pattern limited to colon epithelium. This is consistent with RNA and protein expression data noted in publicly available databases and in the limited number of publications that discuss NXPE1, which describe its importance in gastrointestinal health^42–44^. To facilitate understanding of the NXPE1 patterns, Table S4 provides the results expected from the major hypotheses evaluated, both for proteins and the other assays described later in this study. Tissues with a homozygous T variant of the rs661946 SNP displayed no NXPE1 protein (by IHC) and robust mPAS staining, while heterozygous samples showed NXPE1 immunolabeling but no mPAS staining (Figures 3A and S4, Table S4). Most impressively, some samples with no mPAS staining in the vast majority of crypts contained a crypt or a discrete segment of a crypt that stained robustly, presumably representing focal loss of sialic acid modification. Patients displaying this staining pattern always had a heterozygous (C/T) genotype of the rs661946 SNP, and the rare mPAS staining crypts (less than 1 out of a 1000 crypts) always displayed total loss of NXPE1 protein (Figures 3A, 3B and S4 circled crypts, Table S4). In contrast, NXPE4 and CASD1 staining was independent of mPAS staining (Figure 3B).

**Figure 3:**
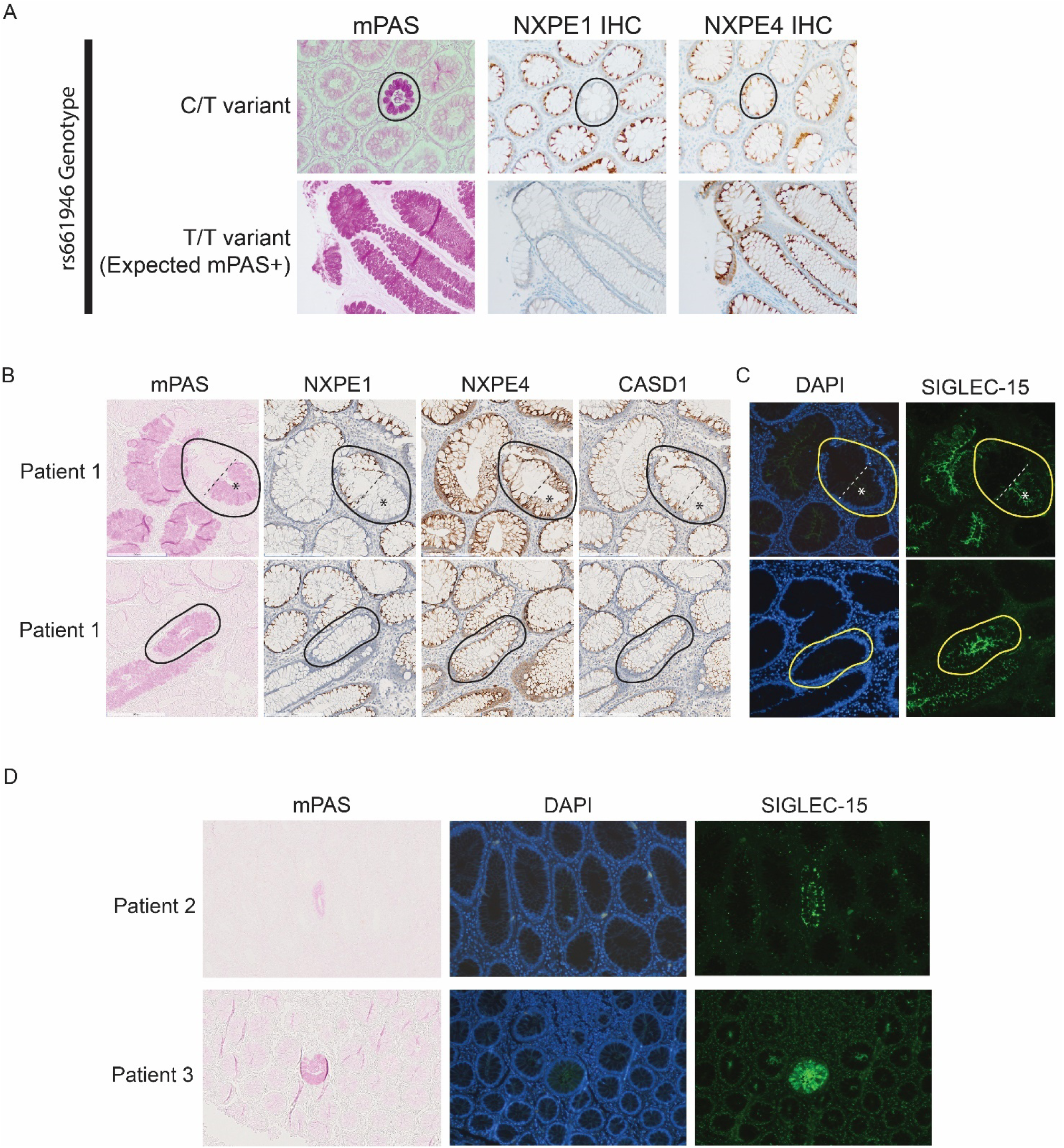
*NXPE1* protein expression is correlated with mPAS and SIGLEC-15 staining. **A)** mPAS, NXPE1 IHC and NXPE4 IHC on adjacent sections from FFPE normal colon tissue with the indicated genotypes for SNP rs661946. Heterozygous samples were primarily negative by mPAS, and positive for NXPE1 and NXPE4, but the images chosen show rare spontaneously mPAS positive crypts. Images are representative examples from more than 20 unique patients, and are shown at 20x. **B)** Adjacent sections of normal colon FFPE tissue heterozygous for rs661946 containing spontaneous mPAS positive crypts stained by IHC for NXPE1, NXPE4 or known sialic acid O-acetyltransferase CASD1. Circles highlight the same crypt in each sample. Note that the crypt in patient 1 denoted with a * is positive only in bottom half of the crypt. NXPE1 protein appears only in the portion of the crypt that is negative for mPAS. Images are representative examples of 10 patients with similar results and are shown at 20x. **C)** Adjacent sections from the same samples in B now stained with SIGLEC-15 and detected by immunofluorescence and DAPI. SIGLEC-15 staining matches the mPAS staining pattern. All images are shown at 20x. **D)** Adjacent sections stained with mPAS or immunofluorescence with SIGLEC-15 on normal FFPE colon tissue heterozygous for the *NXPE1* promoter SNP rs661946. mPAS and SIGLEC-15 stain the same cells/crypts. All images are 20x.

We next sought independent evidence that altered mPAS staining was due to modifications of sialic acid. It is established that the sialic acid-binding immunoglobulin-like lectin (*SIGLEC*) family binds to sialylated glycoproteins^45–48^. O-acetylation modifications are capable of controlling sialic acid-SIGLEC interactions^49,50^. Of twenty SIGLEC/sialic acid binding proteins tested, sialic acid-binding Ig-like lectin 15 (SIGLEC-15) consistently stained colon tissues in the same way as mPAS (Figures 3C and 3D, Table S4). Importantly, SIGLEC-15 staining recapitulated the rare sporadic clusters of NXPE1 negative and mPAS positive crypts in samples heterozygous for the rs661946 SNP (Figures 3C and 3D). SIGLEC-15 is a lectin known to bind Ser/Thr N-acetylgalactosamine (O-GalNAc)-bound sialic acids, and also binds with lower affinity to N-acetylglucosamine (GlcNAc)-bound sialic acids^51,52^. Functionally, SIGLEC-15 has been described as a regulator of osteoclast differentiation as well as a potential immune checkpoint for macrophages^53,54^. SIGLEC-7 also showed some specificity for mPAS positive colon tissue, but the signal was not as consistent or uniform as that of SIGLEC-15 (Figure S5). This observation is supported by previous evidence showing that SIGLEC-7 and SIGLEC-15 bind to similar clusters of O-glycan bound sialic acids^51^.

### Manipulation of NXPE1 protein expression alters sialic acid modification

Using a lentivirus, we introduced a *NXPE1* expression cassette under the control of a Cytomegalovirus promoter into Jurkat cells, derived from a human T-cell leukemia patient. Jurkat was chosen as it expressed sialyl-Tn, a truncated sialic acid-containing O-glycan known to bind to SIGLEC-15 and commonly found in cancer cells, and has very low expression of NXPE1^55^. In a pool of cells infected with a lentivirus containing an NXPE1 open reading frame, increased amounts of NXPE1 protein led to decreased SIGLEC-15 binding by flow cytometry, consistent with the hypothesis that increased *NXPE1* activity would increase the number of modified sialic acids and lower binding of SIGLEC-15 (Figures 4A and S6, Table S4). To confirm these results, we then performed IHC using an anti-sialyl-Tn antibody sensitive to acetylation state. Similar to the lectin-mediated flow cytometry, overexpression of NXPE1 caused robust loss of sialyl-Tn IHC staining on the cell surface of Jurkat cells (Figure 4B).

**Figure 4:**
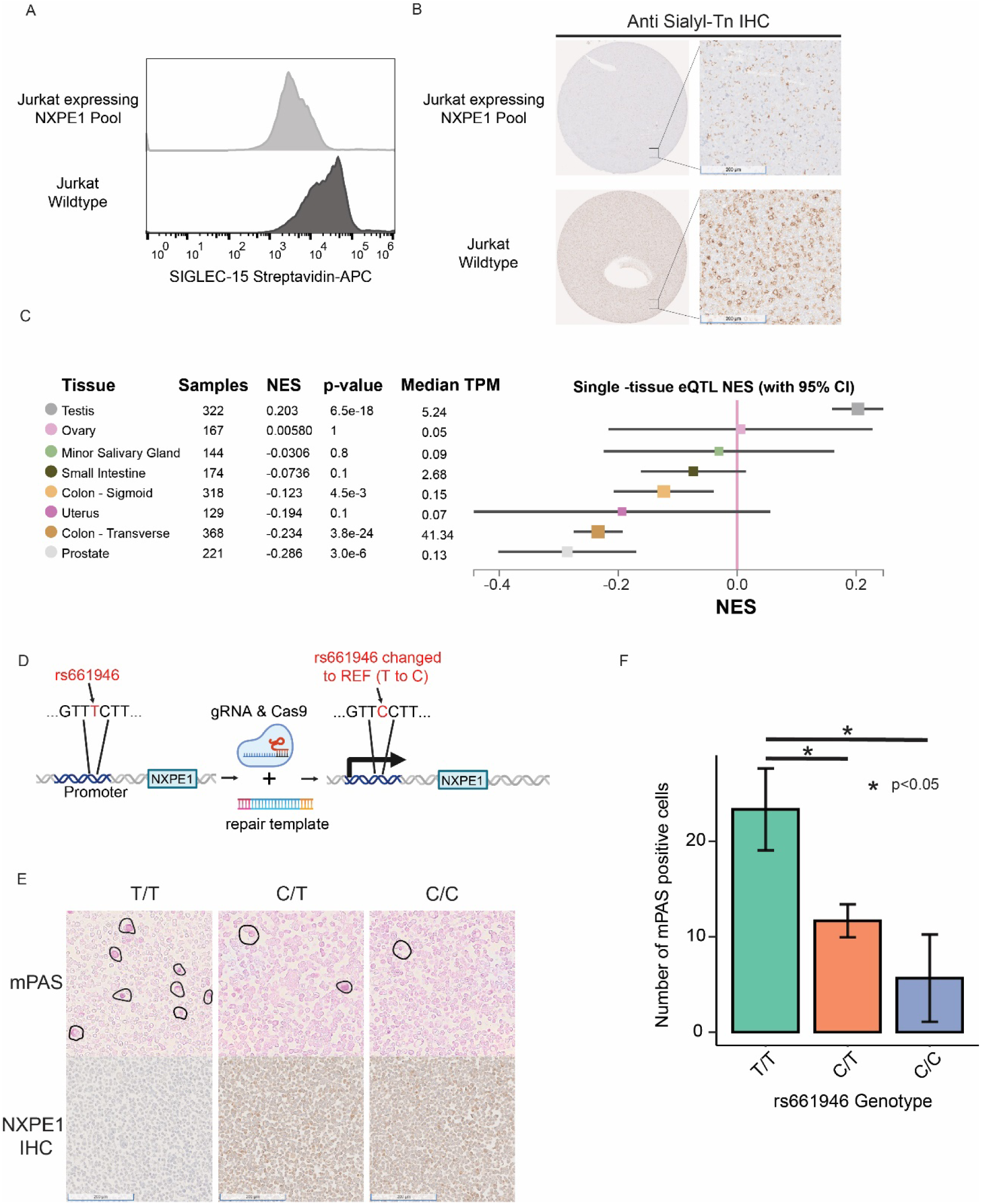
Manipulation of NXPE1 protein expression alters mPAS and SIGLEC-15 staining. **A)** Flow cytometry with biotinylated SIGLEC-15 conjugated to APC on a pooled population of Jurkat cells infected with lentiviral clones containing a NXPE1 expression cassette. **B)** Sialyl-Tn IHC staining on Jurkat wildtype and a pooled population of Jurkat cells overexpressing NXPE1. **C)** GTEx eQTL data for *NXPE1* RNA expression and variant rs661946 by tissue type (see acknowledgements). Tissues not shown in this image had no NXPE1 expression. Normalized Effect Size (NES), which is the same as the beta coefficient (regression slope), is calculated by looking at the effect of the variant allele (T/T) relative to the refences allele (C/C), and negative values indicating less expression from the variant (minor) allele. **D)** Cartoon describing CRISPR-Cas9 knock in approach to change rs661946 in LS180 cells from T/T (homozygous VAR) to C/T (heterozygous) or C/C (homozygous REF). **E)** mPAS and NXPE1 IHC staining on LS180 cells with the indicated genotypes for rs661946, with heterozygous (C/T) and homozygous REF (C/C) being created by CRISPR-Cas9 knock in. Cells positive for mPAS staining are circled. **F)** Counts of the number of mPAS positive LS180 cells with the three possible genotypes for rs661946 as created by knock-in. Quantitation was performed using ImageJ on three different random images from

### *NXPE1* displays differential allelic expression

A review of the literature suggests that expression of NXPE1 protein is largely confined to the colon and rectum^42,56,57^. Consistent with this, the RNA expression levels of *NXPE1* were queried in the GTEx database and found to be highly expressed in normal human colon tissues, averaging 50 transcripts per million^42,57^. We next sought to address the differences in the expression of the two alleles of *NXPE1*. For this purpose, we used SNPs within the transcribed region of *NXPE1* that were in linkage disequilibrium with rs661946. This revealed lower expression of transcripts linked to the T allele in colon-transverse, colon-sigmoid and prostate (Figure 4C and Table S5). Testis also showed an imbalance of allelic expression, but in the opposite direction (more expression from the T, rather than the C allele) (Figure 4C top). Similarly, *NXPE4* also showed fewer transcripts from the variant allele of SNPs in linkage disequilibrium with rs661946 (Table S5). In contrast, the alleles from *RBM7* and *REXO2* in colon-transverse and colon-sigmoid generally showed minimal to no difference in expression (Table S5).

Finally, we wanted to establish whether the SNP rs661946 in the promoter of *NXPE1* was responsible for the inherited changes in protein expression noted earlier in this study. To directly evaluate the effects of changing the T allele to a C allele, colorectal cancer cell lines were surveyed to find one that stained positively with mPAS (Figure S7). We identified one such cell line (LS180), and confirmed the genotype for this cell line as homozygous for the T allele at rs661946, as expected from its mPAS staining (Figure S7, Table S4). CRISPR was used to change the T/T genotype in this line to either the heterozygous C/T or homozygous C/C (Figures 4D and S8). A robust increase in NXPE1 protein and decrease in mPAS staining was observed in the engineered lines with C/T to C/C change (Figures 4E and F).

### NXPE1 transfers acetyl to cytidine-5-monophospho-N-acetylneuraminic acid

We wanted to determine if NXPE1, like CASD1, is able to biochemically transfer an acetyl group from acetyl coenzyme A onto cytidine-5-monophospho-N-acetylneuraminic acid (CMP-Neu5Ac), a modified sialic acid used for biosynthesis of sialic acid derivatives^29^. Co-incubation of the predicted extracellular domain of NXPE1 (AA 60-547) with required co-factors and CMP-Neu5Ac led to the selective formation of Neu5,9Ac_2_, suggesting that NXPE1 mediates addition of acetyl groups to sialic acids at the ninth carbon position (Figures 5A & S9). Expectedly, absence of NXPE1 in the enzymatic mixture prevented biochemical synthesis of O-acetylated sialic acid derivatives (Figure 5A bottom left). Our observations support a model where (1) rs661946 SNP directly modulates the expression of *NXPE1* resulting in variation in NXPE1 activity and (2) NXPE1 mediates sialic acid O-acetylation (Figure 5B).

**Figure 5:**
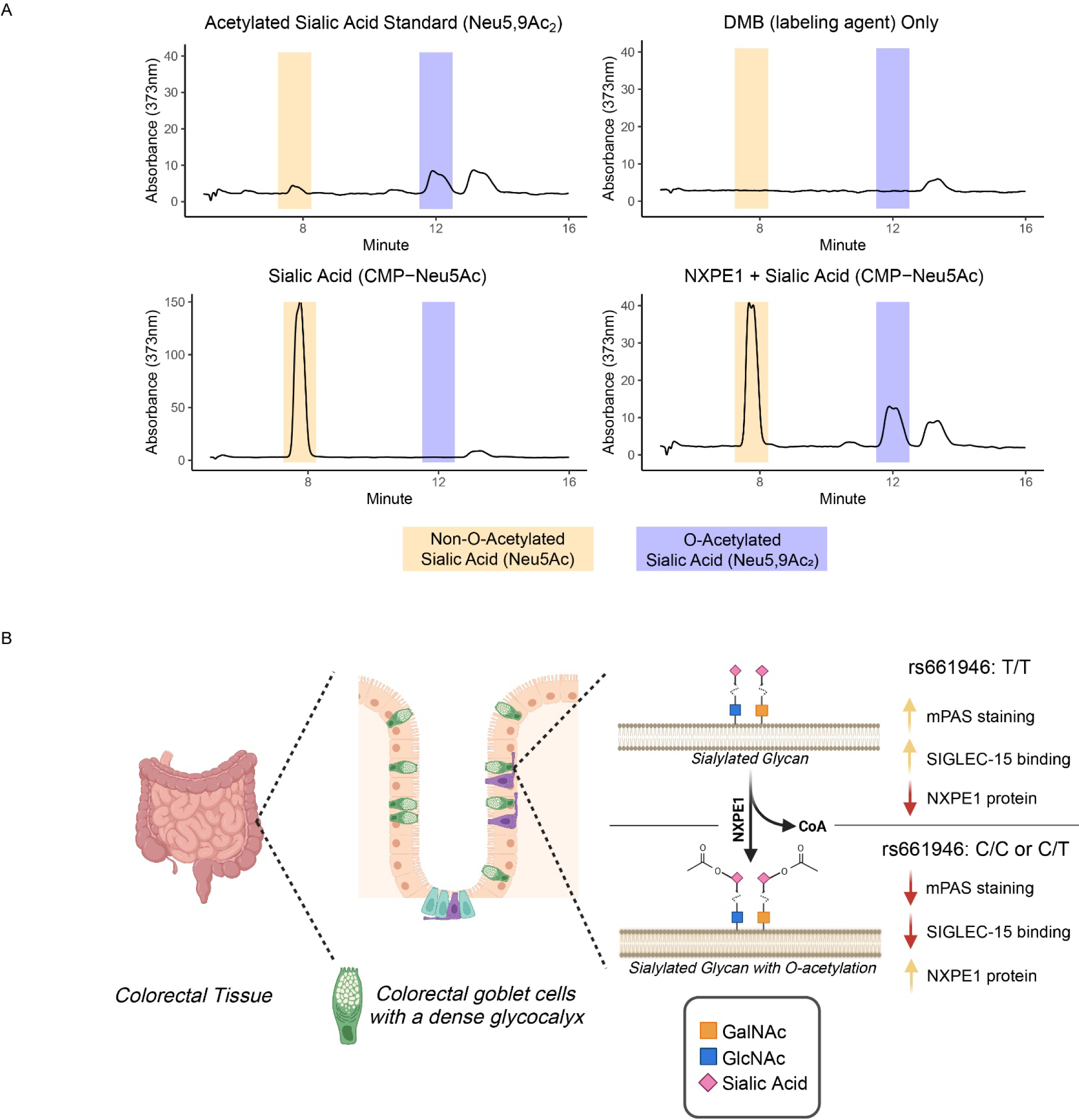
NXPE1 mediates O-acetylation of sialic acids *in vitro*. **A)** DMB-based HPLC readout of an enzymatic reaction containing CMP-Neu5Ac and acetyl coenzyme A, with or without the predicted extracellular domain of NXPE1 (AA 60-547). Note that the O-Acetylated Sialic Acid Standard is composed of ∼77% Neu5,9Ac_2_ (acetylated sialic acid product) and ∼23% Neu5Ac (unacetylated sialic acid). **B)** Cartoon summarizing proposed NXPE1 activity in colorectal tissue. Note that the targeted sialylated glycans shown, GalNAc and GlcNAc, are based on previously published data describing

## Discussion

The search for genes responsible for O-acetylation of sialic acid has been ongoing for several decades, but despite extensive efforts, often using biochemical approaches, only *CASD1* has been described^3,28^. The approach taken in this study was different than those used previously, and our conclusion that *NXPE1* is the acetyltransferase responsible for mPAS staining is based on the following evidence:

(i) Whole genome sequencing of DNA showed that multiple SNPs on chromosome 11q23 were significantly correlated with mPAS staining in colon tissues. No other chromosomal positions were perfectly linked to this phenotype (Figures 1A and B).
(ii) More refined genomic and histochemical studies showed that only a single variant - rs661946, 6 bp upstream of the transcriptional start site of *NXPE1* - was perfectly correlated with mPAS staining in all 112 patients studied. Many other common SNPs in the region, including those within the transcribed region of *NXPE1*, were highly, but not perfectly, correlated (Figure 1D and Table S3).
(iii) *NXPE1* contains motifs identical to those in the catalytic site of the only know human SOAT, as well as viral homologs of SOAT (Figure 2).
(iv) Immunohistochemical studies with antibodies to NXPE1 showed perfect correlations with mPAS staining in the colon. In particular, the rare crypts that were mPAS positive in rs661946 C/T heterozygotes did not express detectable levels of NXPE1 and were immersed in a sea of crypts that did express NXPE1 (Figure 3A).
(v) Of twenty sialic acid-binding proteins in the SIGLEC family tested by immunofluorescence, only SIGLEC-15 consistently stained the same crypts that were stained by mPAS, both in different patients and in different crypts of the same patient. In particular, the rare crypts in rs661946 C/T heterozygotes that bound to SIGLEC-15 were found to be mPAS positive (Figures 3C and D).
(vi) Overexpression of NXPE1 in cell lines resulted in loss of SIGLEC-15 and sialyl-Tn binding (Figure 4A and B).
(vii) In rs661946 C/T heterozygous patients, *NXPE1* transcripts from the alleles containing T were reduced compared to those containing C. GTEx data confirms that there is an allelic imbalance of expression for SNPs located near the *NXPE1* promoter (Figure 4C).
(vii) Knock-in of a C in place of a T allele in a colon cell line resulted in the expected increase in NXPE1 protein and corresponding decrease in mPAS staining (Figure 4E and F).
(ix) NXPE1 protein can acetylate sialic acids at the 9^th^ position *in vitro* (Figure 5A).

While these lines of evidence above strongly implicate *NXPE1* as the SOAT responsible for mPAS staining, many questions remain unanswered. Based on sequence alignments and predicted 3D structures, the catalytic site for *NXPE1* is similar to that of *CASD1.* The specifics of how the acetyl-transfer is performed, and the location on sialic acid targeted by *NXPE1,* remains to be investigated. The preference for SIGLEC-15 to bind NXPE1 modified proteins suggests that these protein targets contain Ser/Thr-GalNac linked sialic acid at the α2-3 or α2-6 position^51,55,58^. However, this remains speculative as we’ve only demonstrated that enzymatically active NXPE1 is capable of acetylating the 9^th^ position of sialic acid, but not the orientation of the sialic acid with the peptide backbone. Furthermore, the *NXPE* family has four members, none of which have been thoroughly characterized, and the potential role of the other three members in modifying the sialoglycome remains speculative.. *In silico* 3D structures, however, predict that all of the family members contain a conserved GDS and HPP motif exposed to solvent. This leads us to speculate whether they may also perform similar sialic acid acetylation activity in other tissues, on different positions of sialic acid, or on sialic acids with different linkages^36^.

There are potential practical implications of our findings. Genome-wide association studies have shown that the genomic region containing *NXPE1* is linked to inflammatory bowel disease^43,59^. It is possible that *NXPE1*, or the closely positioned *NXPE4*, are responsible for this linkage^60^. If so, then one could imagine that NXPE enzymatic inhibitors could be used to treat a subset of patients with this disease. Second, loss of 11q23 is found in more than 20% of colorectal cancers^61,62^. Approximately 50% of these patients will be C/T heterozygotes at rs661946. When the C allele is lost in the cancer cells of these patients, they are no longer O-acetylated on the sialic acids recognized by mPAS or SIGLEC 15. This creates potential opportunities for specifically targeting the non-O-acetylated cancer cells while sparing healthy O-acetylated cells. With potential to impact health and wellbeing in a number of ways, there is clearly a lot left to explore in this exciting field.

## Data Availability

All data produced are available in the manuscript or online at the European Genome-phenome Archive under accession number EGAS00001007704

https://ega-archive.org/

## Acknowledgements

The authors would like to thank Dr. Sujayita Roy, Sarah Hughes and Dr. Alan Meeker of the Johns Hopkins Sidney Kimmel Comprehensive Cancer Center Oncology Tissue Services Core for their assistance with IHC staining and optimization. They would also like to thank Dr. Tatianna Larman and Dr. Maged Zeineldin for their help with reviewing the manuscript. The Genotype-Tissue Expression (GTEx) Project was supported by the Common Fund of the Office of the Director of the National Institutes of Health, and by NCI, NHGRI, NHLBI, NIDA, NIMH, and NINDS. The data used for the analyses described in this manuscript were accessed from the GTEx Portal on 11/2023. Tissue expression information was also accessed from the Human Protein Atlas proteinatlas.org on 11/2023. This work was supported by Oncology Core CA 06973 (B.V., K.W.K., N.P.); The Virginia and D.K. Ludwig Fund for Cancer Research (B.V., K.W.K., N.P., C. B.); The Sol Goldman Sequencing Facility at Johns Hopkins (B.V.); NIH Cancer Center Support Grant P30 CA006973; Lustgarten Foundation for Pancreatic Cancer Research (B.V., N.P., K.W.K., and S.Z.); The Commonwealth Fund (B.V., N.P., K.W.K., S.Z., and C.B.); Howard Hughes Medical Institute (B.V.).

## Supplemental Tables

S1-Whole genome sequencing statistical summary

S2-List of interesting SNPs from WGS analysis

S3-Targeted sequencing genotypes and mPAS staining

S4-Staining reference

S5-GTEx results from NXPE1 and NXPE4 eQTL

## Extended Data

Figures S1 to S9

## Materials and Correspondence

Correspondence or request for materials should be directed to Kenneth Kinzler Ph.D. or Nicolas Wyhs Ph.D.

## Author Contributions

NW, KK and BV conceptualized the study. SS, SZ, BV, NP, CB, KG, PB and SK provided materials and advice on study and experiment design. NW, BSL, AC, LD, MP, BV, KK and KG performed experiments. NW, KK and BSL wrote the manuscript.

## Methods

### Mammalian cell lines and cell culture

Jurkat, HEK293T, and LS180 cells were purchased from The American Type Culture Collection (Virginia, USA). Jurkat cells (ATCC, Cat #TIB-152) are male in origin and were grown in RPMI 1640 Medium (ATCC, Cat #30-2001), supplemented with 10% fetal bovine serum (HyClone, Utah, USA, Cat #16777-006) and 1% Penicillin-Streptomycin (Gibco, USA, Cat #15140122). LS180 cells (ATCC, Cat #CL-187) are female in origin and were grown in EMEM (ATCC, Cat #30-2003), supplemented with 10% FBS (HyClone, Utah, USA, Cat #16777-006) and 1% Penicillin-Streptomycin (Gibco, USA, Cat #15140122). HEK-293T cells (ATCC, #CRL-3216) were grown in DMEM (Gibco, USA # 11965092) supplemented with 10% FBS (HyClone, Utah, USA, Cat #16777-006) and 1% Penicillin-Streptomycin (Gibco, USA, Cat #15140122). In vitro cells were maintained at 37°C with 5% CO2. Mycoplasma testing was performed by The Genetic Resources Core Facility at Johns Hopkins School of Medicine (Maryland, USA).

### Whole genome sequencing (WGS) workflow

21 samples: 8 mPAS positive, 8 mPAS negative, and 5 mPAS negative with rare positive crypts - were selected for whole genome sequencing. Matched FFPE sections and plasma pellets were commercially obtained from ILSbio LLC (Chestertown MD) for each sample. DNA was purified from on average 5mL of plasma using a DNeasy Blood & Tissue Kit (Qiagen 69504), and quantified by Nanodrop 2000 (ThermoFisher Scientific). Library preparation was performed using a ThruPLEX® DNA-Seq kit (Takara R400675) per manufacturer’s instructions. Each sample was sequenced on 3 or 4 lanes of a Hiseq 4000 (Illumina) with paired end 100 cycle settings per manufacturer’s instructions. Fastq files for each sample are publicly available at the European Genome-Phenome Archive (https://ega-archive.org/) under accession number EGAS00001007704. VCF files were created for each using the cancer genomics cloud (Seven Bridges)^63^. Briefly, alignment and VCF generation was performed with Whole Genome Analysis - BWA + GATK 2.3.9-Lite^64,65^. VCF files were then moved into SQL for further analysis including genotype assignment and comparison. SNPs were considered a perfect genotype-phenotype match if one of two conditions were met: 1) all positive mPAS staining samples contained an alternate (VAR) genotype, negative staining with rare positive contained a heterozygous genotype, and negative staining contained either a heterozygous or homozygous reference genotype. 2) All mPAS positive staining samples contained a reference genotype, negative mPAS staining with rare positive contained a heterozygous genotype, and negative mPAS staining samples contained a heterozygous or homozygous alternate (VAR) genotype. Data was also analyzed in a less stringent form, allowing for 1-2 mismatches between genotype and phenotype. Variant consequence analysis was performed using Variant Effect Scoring Tool (https://www.cravat.us/CRAVAT/)^66^. Plot of perfectly concordant SNPs (Figure S1) was performed using the package mapsnp in R^67^. GWAS and association case-control analysis was performed on 10,421,023 variants using PLINK v2.00a6 64-bit with the following settings: gender set as a covariate, eliminated SNPs with a minor allele frequency below 0.1, assumed dominant genotype, only considered biallelic SNPs, and removed SNPs not in Hardy Weinberg Equilibrium. Linkage disequilibrium (LD) and haplotype block analysis was performed using PLINK v1.90b7.2 64-bit (11 Dec 2023), pairwise LD comparisons were made between all SNPs within 1Mb of each other on chromosome 8 and 11. GWAS study plots generated with the R packages qqman v0.1.9 (http://pngu.mgh.harvard.edu/purcell/plink/)^68,69^, GWLD v1.3.6^70^ and Ldheatmap v1.0-5.

### Targeted sequencing workflow

Targeted sequencing validation was performed on DNA from normal colon or normal adjacent tumor FFPE samples obtained commercially from ILSbio LLC (Chestertown, MD). Six 5µm sections were pooled together into a single tube for each of the 91 samples. DNA was extracted using a QIAamp DNA FFPE Tissue Kit (Qiagen 56404). DNA was quantitated using nanodrop 2000 (ThermoFisher Scientific). PCR was performed on target locations using Phusion Flash High-Fidelity PCR Master Mix (ThermoFisher Scientific F548L) following manufacturer’s instruction and primers listed below. Sanger sequencing of the product was performed by Genewiz.

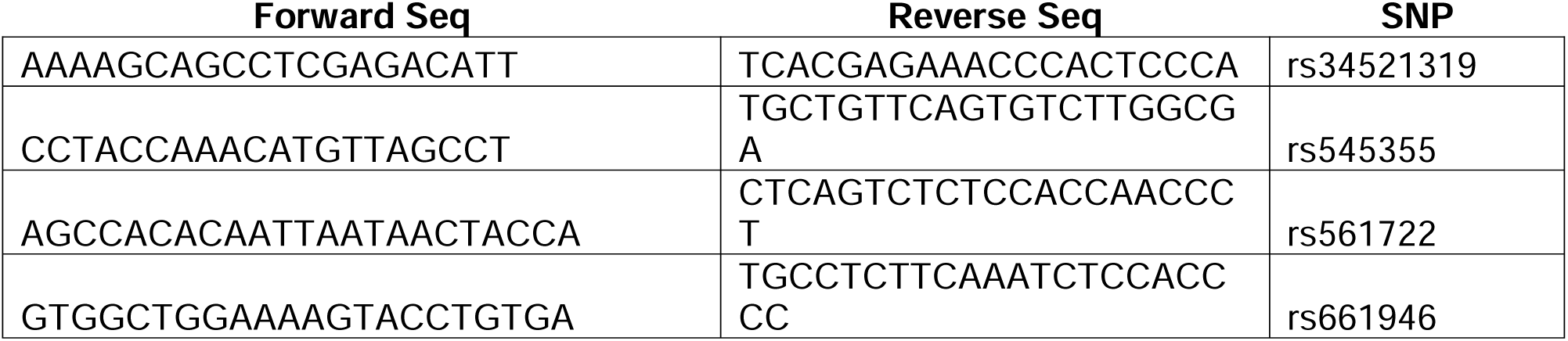

### Multiple Sequence Alignment and in-silico 3D structure prediction

Multiple sequence alignment was done using NCBI-BLASTp, COBALT and Jalview^71–73^. 3D structure prediction was accomplished using AlphaFold^40^. All were accessed 11/2023.

### Overexpression in cell lines

Overexpression cassettes (pLenti-C-mGFP-P2A-Puro Vector #RC206474L4 from Origene) were transfected into HEK-293T cells with MegaTran 2.0 (Origene TT210002) with psPAX2 (Addgene #12260) and pMD2.G (Addgene #12259) in a 10:3:1 ratio (30µg of total plasmid DNA per well) respectively following manufacturer’s instructions. Lentivirus was harvested after 48hrs, and a second time at 72hrs. Harvests were combined, run through a 0.45 PES filter (Millipore-SLHPM33RS), and concentrated with Lenti-X concentrator (Takara #631232). Virus stocks were stored in 1mL of DMEM (Gibco, USA # 11965092) with 10% FBS (HyClone, USA, Cat #16777-006) and concentrations determined by p24 ELISA (Takara #632200). Non-treated 48 well plates were treated with RetroNectin, according to manufacturer’s instructions (Takara #T100). Using RetroNectin-treated plates, 50,000 target cells were transduced by spinoculation at 800xg for 2hrs at 37°C with 300µL virus. Cells were washed and then monitored for GFP and target gene expression. Selection for 14 days was performed with 1µg/mL of puromycin (InvivoGen ant-pr-1).

### CRISPR KI of cell lines

gRNA was obtained from Integrated DNA Technologies (IDT) (#CD.HC9.KDJS8530.AA), repair template was also obtained from IDT (#CD.HC9.QPVN2588). gRNA complex was performed by mixing gRNA (50µM final concentration) and Alt-R® CRISPR-Cas9 tracrRNA (IDT #1072533)(50µM final concentration), heating to 95°C for 5min then allowing to cool to room temperature on benchtop. RNP complex was performed by mixing 3µL of the above gRNA complex with 2.4 µL of Alt-R™ S.p. Cas9-GFP V3 (IDT #10008161) (final concentration 30µM for each) and incubating at room temperature for 15min. LS180 cells were trypsinized (Gibco # 25200056), pelleted and washed twice with PBS (Gibco # 10010023), then suspended in SG Nucleofection buffer (Lonza, #V4XC-3024) at 25e6cells/mL. A total of 5e5 cells were electroporated by mixing 5µL of RNP complex from above, 1.2µL of HDR Donor Oligo, 1.2µL of Alt-R™ HDR Enhancer V2 (IDT #10007910), 20µL of the LS180 cell suspension in SG buffer (see above), and 0.1µL of PBS (all reagents final concentration of 5µM). Cells were immediately electroporated on a 4D Nucleofector X-Unit (Lonza, AAF-1003X) using pulse code DS-150A. Cells were then removed and plated into complete media and incubated for 5 days. Once enough cells were present, cells were clonally selected by limited dilution as previously described^74^. Accurate knock in was confirmed by PCR using Phusion Flash High-Fidelity PCR Master Mix (ThermoFisher #Scientific F548L) following manufacturer’s instruction and primers listed below. Sanger sequencing of the product was performed by Genewiz.

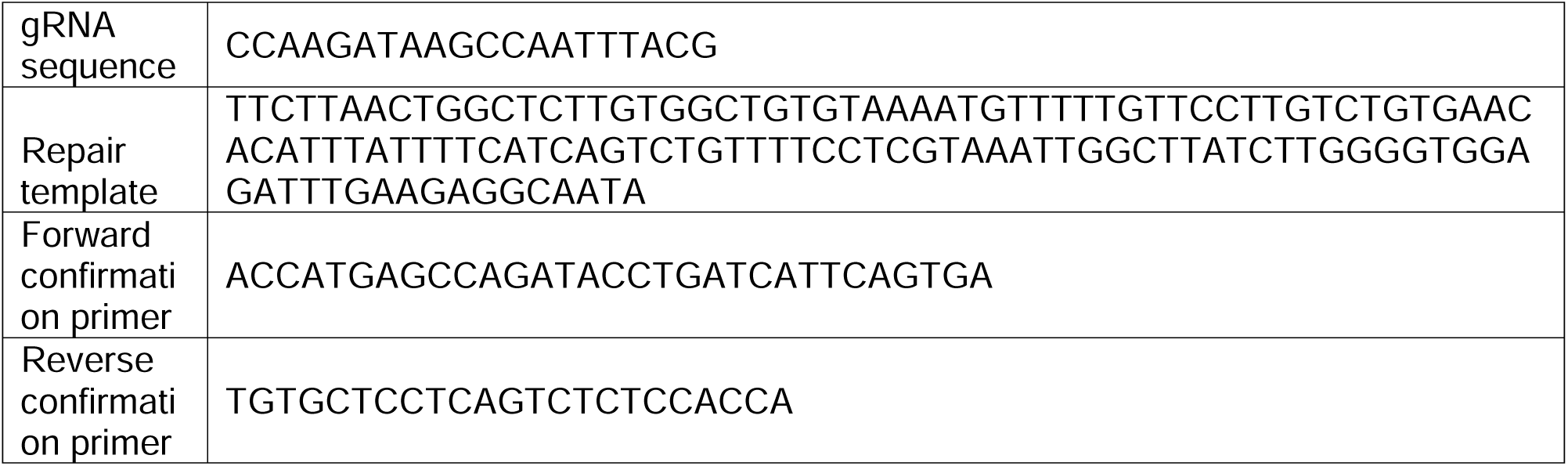

### Immunohistochemistry (IHC) analysis

Immunostaining was performed at the Oncology Tissue Services Core of Johns Hopkins University School of Medicine. Immunolabeling was performed on formalin-fixed, paraffin embedded sections on a Ventana Discovery Ultra autostainer (Roche Diagnostics). Briefly, following dewaxing and rehydration on board, epitope retrieval was performed using Ventana Ultra CC1 buffer (Roche Diagnostics, Cat #6414575001,) at 96°C for 64 minutes. Primary antibody, anti-CASD1 (1:100 dilution; Invitrogen, Cat #PA5-60700, Lot #Xi3700359), anti-NXPE4 (1:1000 dilution; Sigma-Aldrich, Cat #HPA042801, Lot #R39941), anti-NXPE1 (1:200 dilution; Santa Cruz Biotechnology, Cat #sc-514349,) or Anti-Sialyl Tn antibody [STn 219] (1:50 dilution; Cat #ab115957, Lot 1063860-1, Abcam) diluted in Antibody Diluent with Casein (Roche Diagnostics, Cat #6440002001,); was applied at 36°C for 60 minutes. Primary antibodies were detected using an anti-rabbit HQ detection system (Roche Diagnostics, Cat #7017936001 and 7017812001,) followed by Chromomap DAB IHC detection kit (Roche Diagnostics, Cat #5266645001), counterstaining with Mayer’s hematoxylin, dehydration and mounting.

### Mild periodic acid Schiff staining

Staining for mPAS was performed by the Johns Hopkins Reference Pathology Lab. Briefly, slides were dewaxed and hydrated in distilled water. Slides were then washed in 0.1M acetate buffer, pH 5.5 (Thermo Fisher, Cat #AM9740), at 2°C for five minutes. Slides were subsequently treated with 1 mM (0.02%) NaIO_4_ (Sigma Aldrich, Cat #P7875-100G) in 0.1M acetate buffer, pH 5.5, at 4°C for 2min, and then washed in 1 % aqueous glycerol (Sigma Aldrich, Cat #G5516) for five minutes. Slides were washed in distilled water for five minutes and treated with Schiff’s reagent (Sigma Aldrich, Cat #3952016-500ML) at room temperature for 15 minutes. Slides were washed three times in 0.5% K_2_S_2_0_5_ (Sigma Aldrich, Cat #60508) in 0.05M hydrochloric acid (Sigma Aldrich, Cat #2104-50ML) for five minutes, and then washed in running tap water for five minutes. Slides were then washed in distilled water for five minutes. Finally, slides were dehydrated, cleared, and mounted with a coverslip.

### Immunofluorescence (IF) analysis

Blocking was performed by incubating paraffin-embedded tissue samples using 1% BSA with 2% FBS in PBS-T, for a total of 30 minutes. Immunostaining was performed by incubating samples with both primary and secondary antibodies overnight at 4°C using the following reagents: recombinant human Siglec-15-Fc (5µg/mL; R&D Systems, USA, Cat #9227-SL-050) and Anti-Human IgG Alexa 488 (3µg/mL; Jackson ImmunoResearch, USA, Cat # 109-545-170). Samples were thoroughly washed with PBS and mounted in Prolong Gold Antifade Reagent with 4’, 6-diamidino-2-phenylindole, dihydrochloride (DAPI). Other SIGLEC constructs tested were also obtained from R&D Systems and used at an equivalent concentration.

### Cell staining and flow cytometry

Jurkat cells with NXPE1 overexpression were suspended at 1×10^6^ cells/mL in staining buffer and incubated with constructs at relevant concentrations for 30 minutes on ice, in the dark. Primary staining was performed with SIGLEC-15 (Acro Biosystems, Cat #SG5-H82E9) monomer at a concentration of 5µg/mL. Secondary staining was performed with APC-conjugated streptavidin (BioLegend, Cat #405207) at a concentration of 2µg/mL. Single, live cells were isolated using forward and side scatter characteristics. Flow cytometry data was analyzed using FlowJo v. 10.1 software.

### NXPE1 expression plasmid and protein production

Expression construct and protein expression was performed by GeneArt (ThermoFisher Scientific). Briefly, expression constructs for the predicted extracellular domain of NXPE1 (AA 60-547) were synthesized as human codon-optimized gene fragments and cloned into a pcDNA3.4-TOPO vector backbone with an IL2 secretion signal followed by a 6x histidine tag and GGGG linker, then NXPE1 protein sequence. NXPE1 expression plasmid DNA was purified from transformed DH10B E.coli competent cells through GeneArt (ThermoFisher). Recombinant NXPE1 was expressed by GeneArt (ThermoFisher) in Expi293 cells and purified using the HisTrap column (GE29-0510-21, Millipore Sigma). Recombinant NXPE1 was stored in PBS with 10mM Beta mercaptoethanol (M6250, Sigma-Aldrich).

### *In vitro* NXPE1 biochemical assay using DMB-based HPLC

Enzymatic assays to determine sialic acid O-acetyltransferase activity *in vitro* were adapted from^29^. 50mM MES pH 6.5 (Thermo Scientific Chemicals, Cat #J61587.AK), 10mM MnCl_2_ (Sigma-Aldrich, Cat #M1787), 1mM acetyl-CoA (Sigma-Aldrich, Cat #A2056), and 1.25mM CMP-Neu5Ac (Sigma-Aldrich, Cat #5.05223) were incubated with or without 5μg recombinant NXPE1 (aa 60-547) in a 20μL reaction volume for 3h at 37°C. Sialic acids were then released via acid hydrolysis by adding 2M propionic acid (Sigma-Aldrich, Cat #402907) to each sample for 1h at 80°C. Released sialic acid samples were subsequently labelled with 4,5-methylenedioxy-1,2-phenylenediamine dihydrochloride (DMB) following manufacturer’s instructions (Agilent, Cat #GKK-407). DMB-labelled samples,reference panel, Neu5Ac standard (Nacalai, Cat #08371-36), Neu5Gc standard (Ludger/QA-Bio, Cat #CM-NEU-GC-01), and Neu5,9Ac_2_ standard (Ludger/QA-Bio, Cat #CM-NEU5,9AC2-01) were analyzed on a GlykoSep R HPLC column (Agilent) via an Agilent 1260 Infinity HPLC system fitted with a 1260 Infinity Multiple Wavelength Detector. Isocratic elution with acetonitrile/methanol/water (9:7:84, v/v) was performed at a flow rate of 0.7mL/min and absorbance was measured at the excitation wavelength of DMB (373nm).

### Data Availability

Fastq files for all 21 samples that underwent whole genome sequencing are publicly available at the European Genome-Phenome Archive (https://ega-archive.org/) under accession number EGAS00001007704.

### Statistics

Statistics were performed using R version 4.3.2 and RStudio (RStudio 2023.09.0+463 “Desert Sunflower” Release (b51c81cc303d4b52b010767e5b30438beb904641, 2023-09-25). GWAS and association case-control p value analysis was performed using PLINK v2.00a6 64-bit, and linkage disequilibrium r^2^ values obtained using PLINK v1.90b7.2 64-bit (11 Dec 2023) (http://pngu.mgh.harvard.edu/purcell/plink/)^68^. Hardy-Weinberg equilibrium by Haldane’s exact test performed using R-package HardyWeinberg (version 1.7.5)^75^. Fisher’s Exact Test and student T-test were performed using base R-package stats. Multiple sequence alignment expect values were calculated by the BLASTp software and listed here as reported^73^. pLDDT values were calculated by AlphaFold during modeling and listed as reported. Quantification of knock-in mPAS stained slides was performed using ImageJ and student T-test in Excel^76^.

---

**Figure S1:**
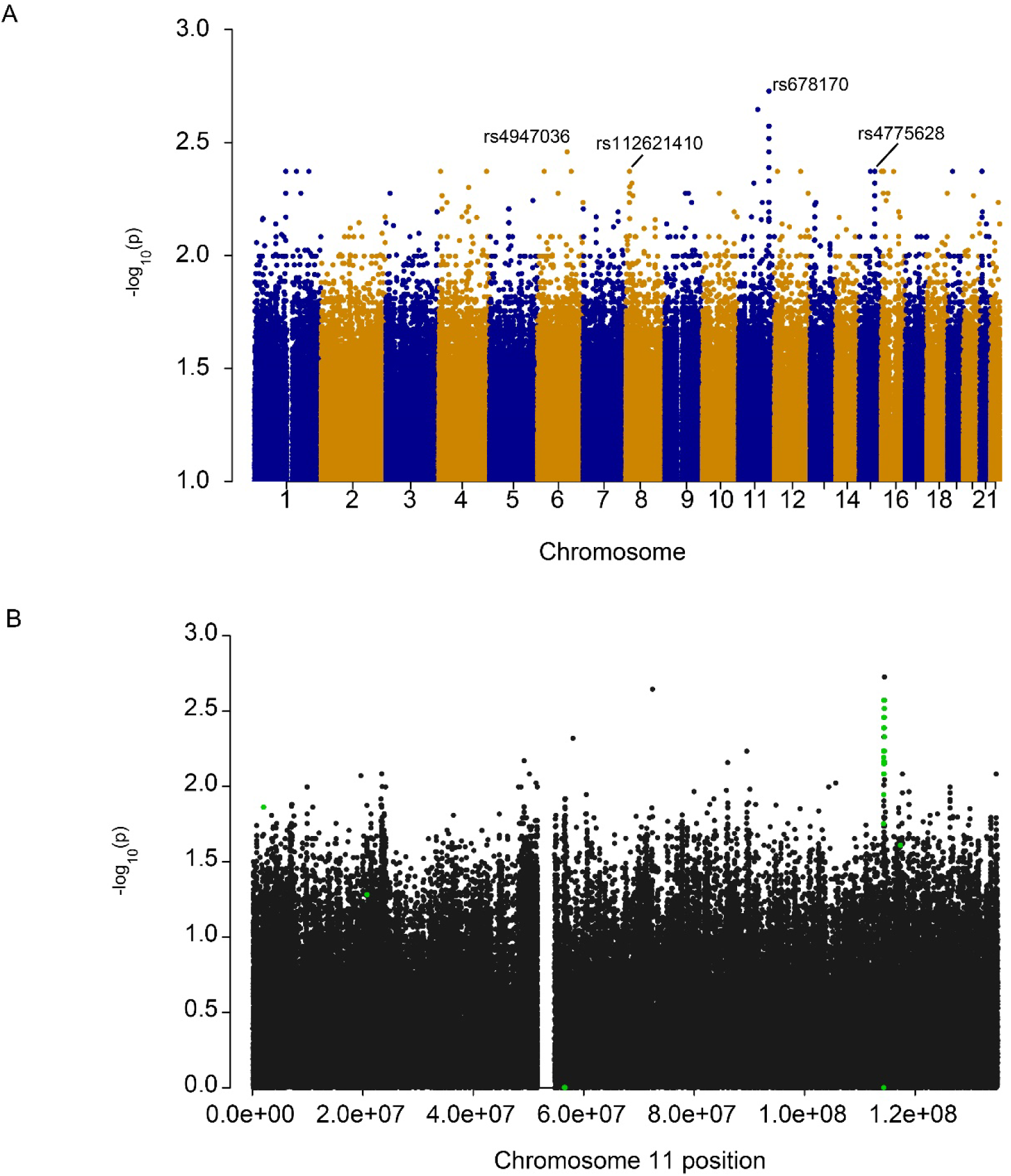
A) Manhattan plot showing SNP association with normal colon mPAS staining genome-wide. B) Same as A but showing only chromosome 11. SNPs with perfect genotype-phenotype concordance are highlighted in green, and cluster in a suspected haplotype near 11q23.2.

**Figure S2:**
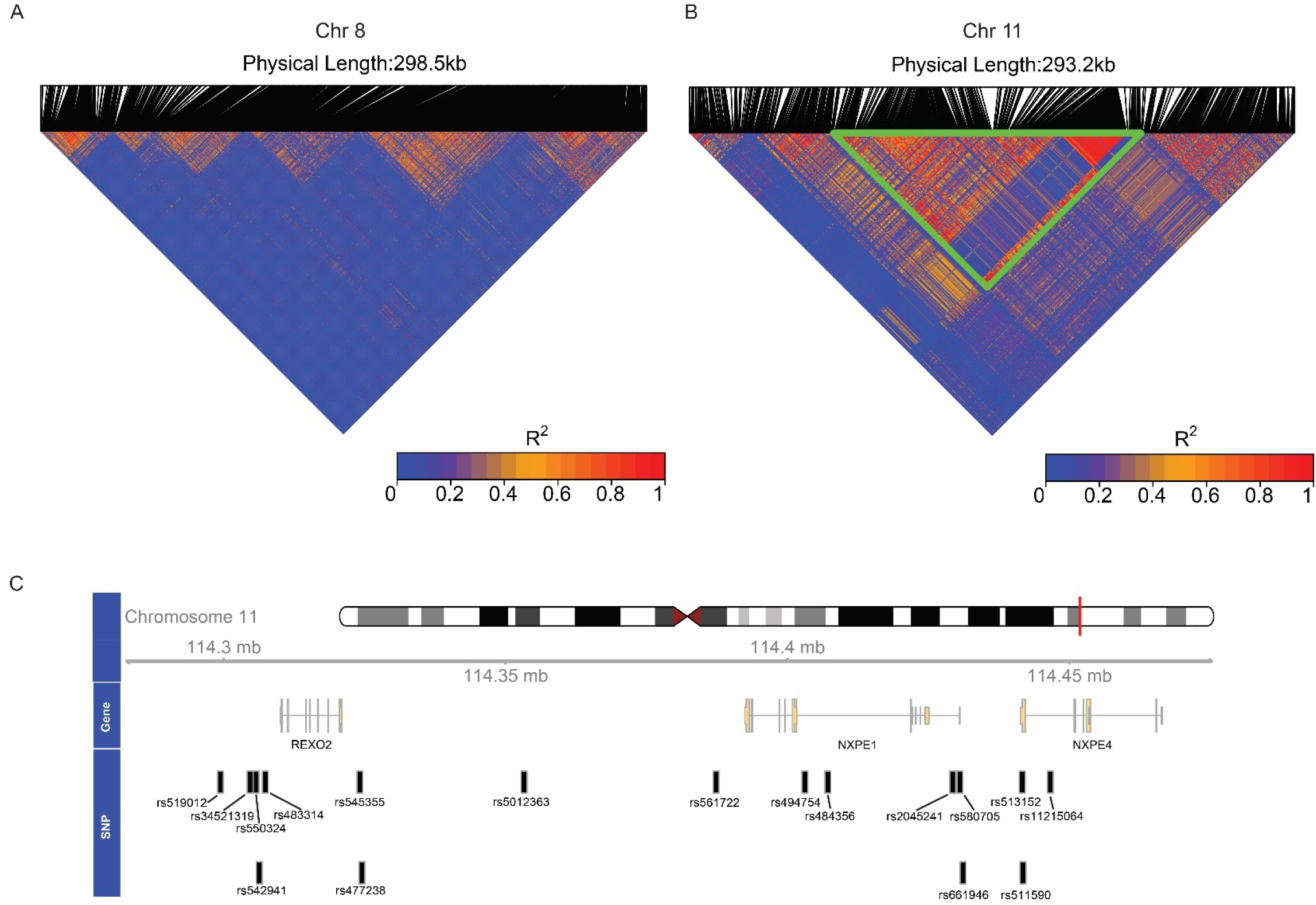
**A)** Linkage disequilibrium plotted as r^2^ values for 300kb surrounding the most significant SNP identified in the genome wide association study of mPAS staining near SNP rs112621410 on chromosome 8 and **B)** rs678170 on chromosome 11. The green highlighted area indicates the region with SNPs demonstrating a perfect concordance between genotype and mPAS phenotype. **C)** Map showing a zoomed in view of the green highlighted region in B and noting the location of SNPs with perfect concordance between genotype and mPAS staining phenotype. Note that the gene RBM7 is also within the discovered haplotype, but contains no perfectly matching SNPs and thus does not appear in this figure.

**Figure S3:**
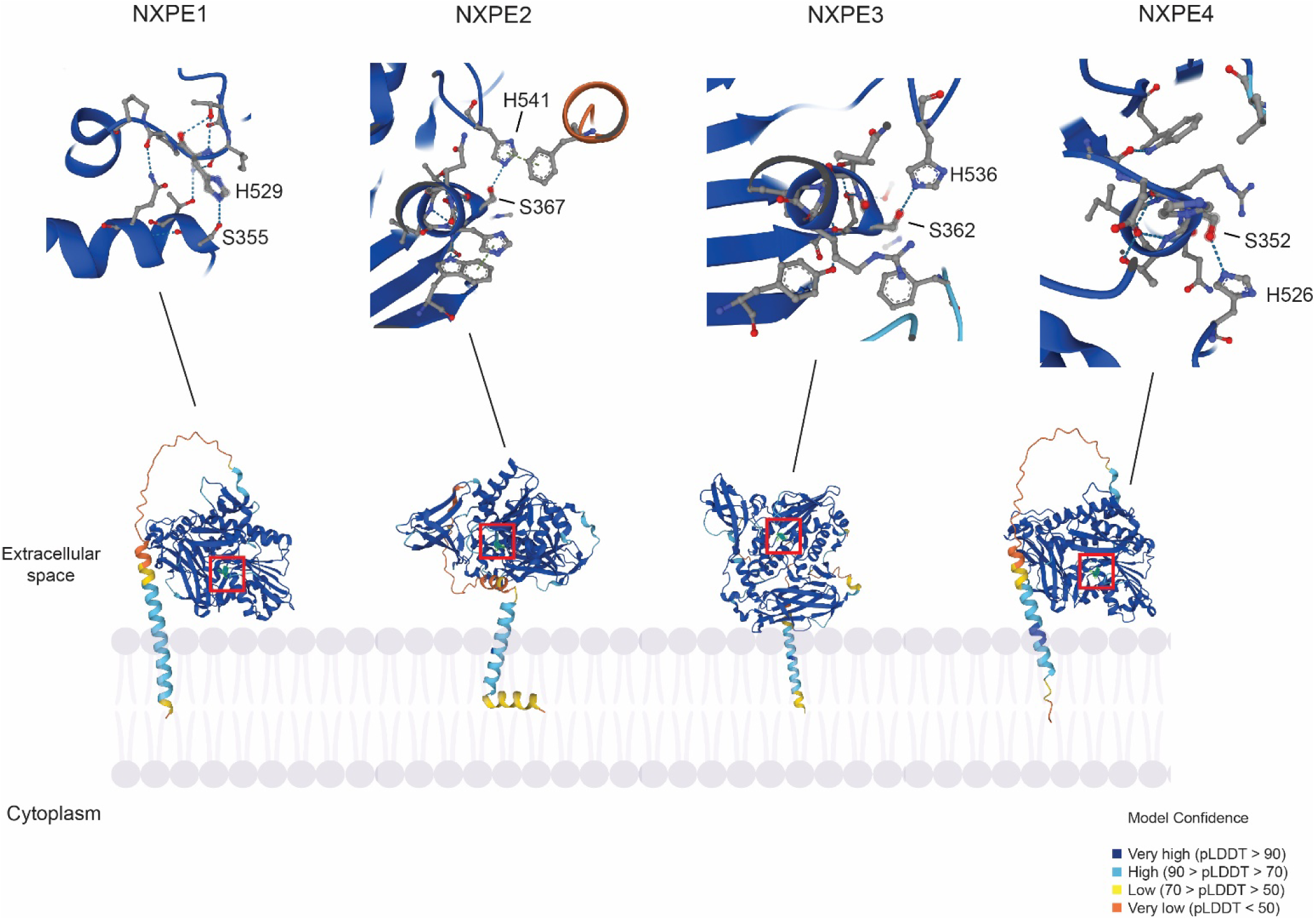
*In silico* predicted 3D structures for NXPE family members NXPE1 (Q8N323.2), NXPE2 (Q96DL1.2), NXPE3 (Q969Y0.1) and NXPE4 (Q6UWF7.1). Active site serines and their corresponding histidines are shown (top). All family members are predicted to contain a single transmembrane alpha helix, with the majority of protein in the extracellular (intra-Golgi/ER) space, consistent with glycan location inside and outside the cell. All 4 members contain a conserved Gly-Asp-Ser (GDS) motif, which is also present in known sialic acid acetyltransferase active sites(Figure 2A), with the serine predicted to collocate near a histidine (top). This figure redisplays NXPE1 structural data from Figure 2D. All structures shown in this figure are predictions based on AlphaFold accessed 11/2023.

**Figure S4:**
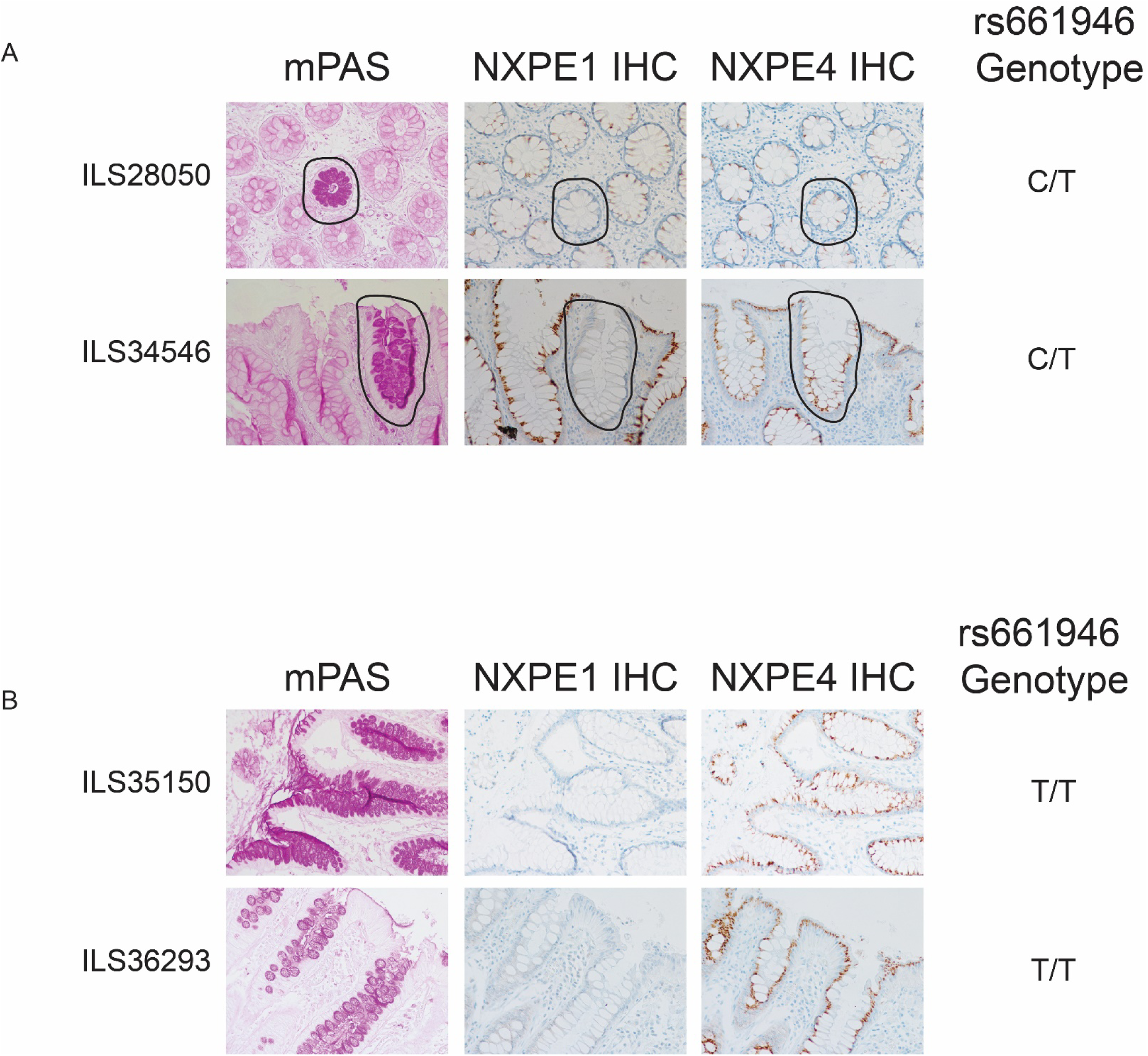
A) mPAS, NXPE1 IHC and NXPE4 IHC staining on FFPE normal colon tissue with heterozygous (C/T) and B) homozygous-VAR (T/T) genotypes for SNP rs661946. Heterozygous samples were primarily negative by mPAS, and positive for NXPE1 and NXPE4, but the images chosen show rare cases of spontaneously mPAS positive crypts (circled). Images are shown at 10x. This figure provides additional images supporting figure 3A.

**Figure S5:**
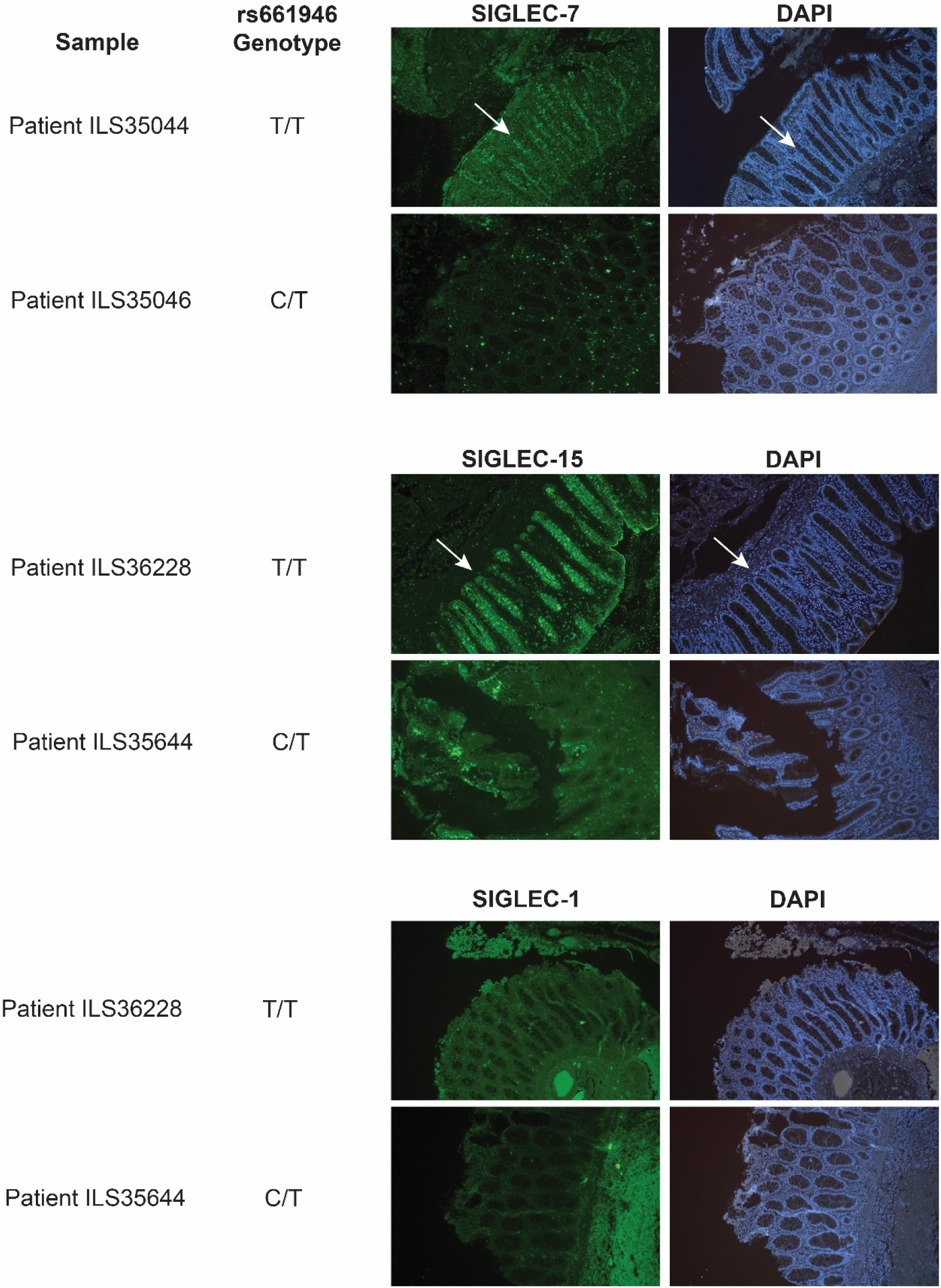
SIGLEC-7 (top), SIGLEC-15 (middle) and SIGLEC-1 (bottom) IF staining on normal FFPE colon tissue. The homozygous VAR (T/T) genotype is expected to stain positive for lectin. Arrows show examples of specific staining in goblet cells. SIGLEC-7 displays specific staining of crypt goblet cells, but not as robustly as SIGLEC-15. SIGLEC-1 is shown as a control and does not stain.

**Figure S6:**
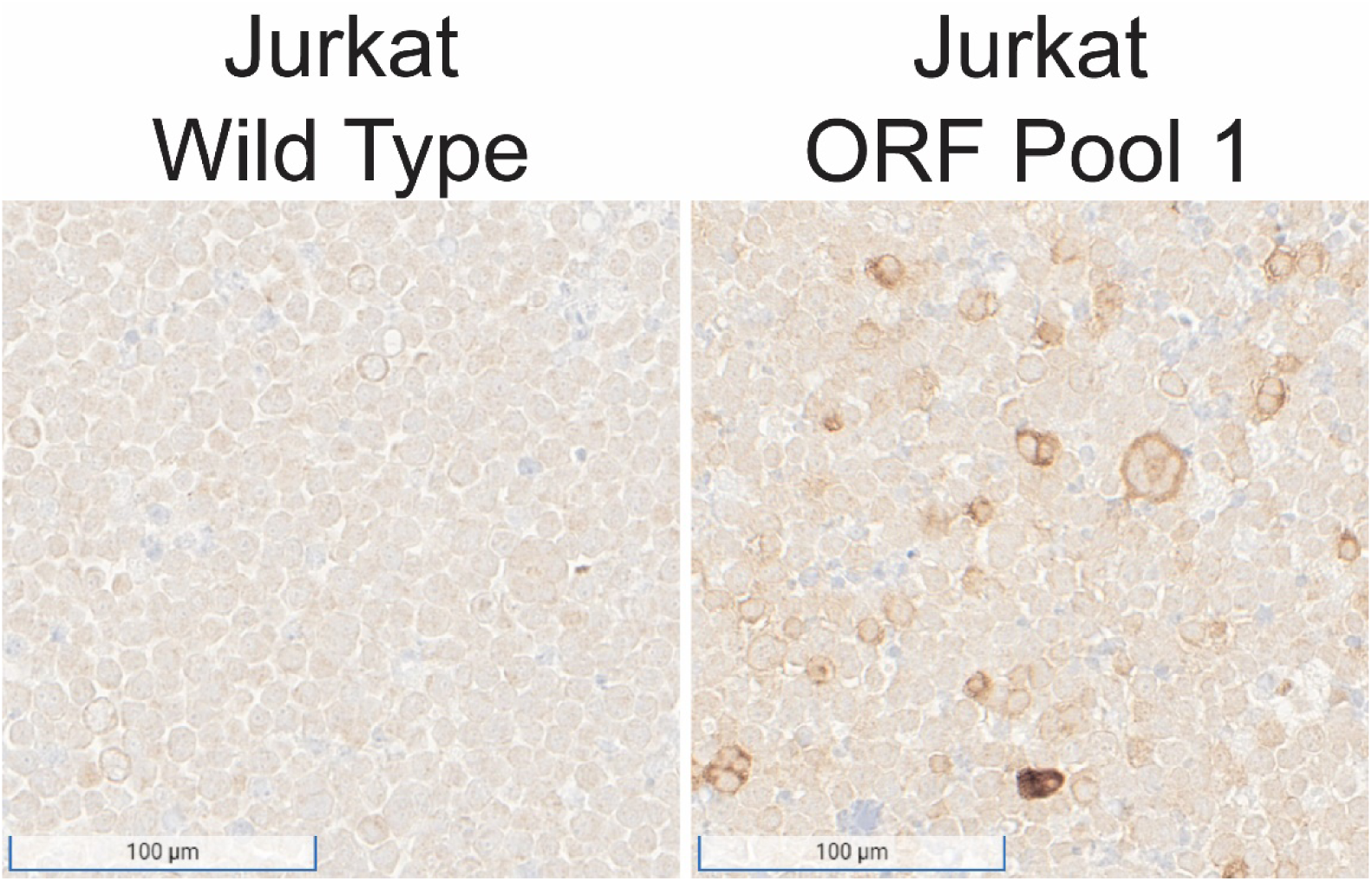
IHC of NXPE1 protein on Jurkat parent cells and a pool of Jurkat cells transfected with an NXPE1 open reading frame.

**Figure S7:**
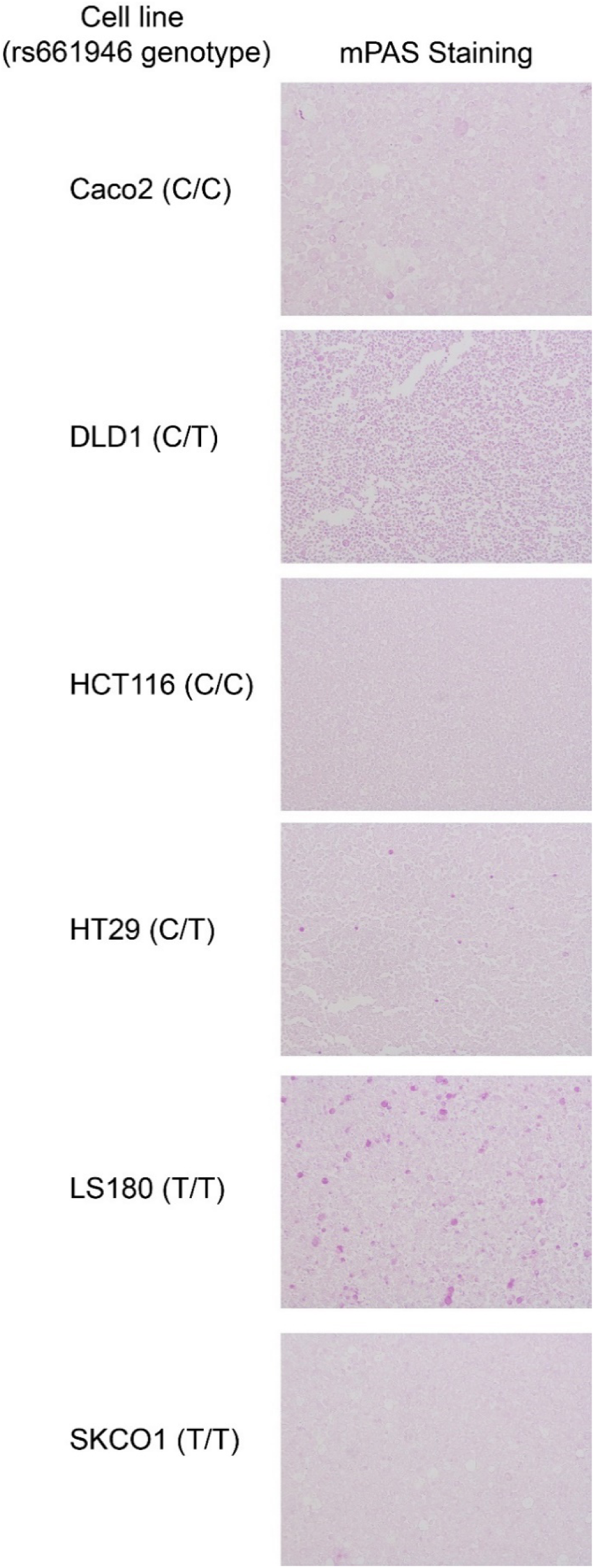
mPAS staining on various colon cancer cell lines. SNP rs661946 genotype is noted in parenthesis next to cell line name, T/T genotype is expected to stain positive.

**Figure S8:**
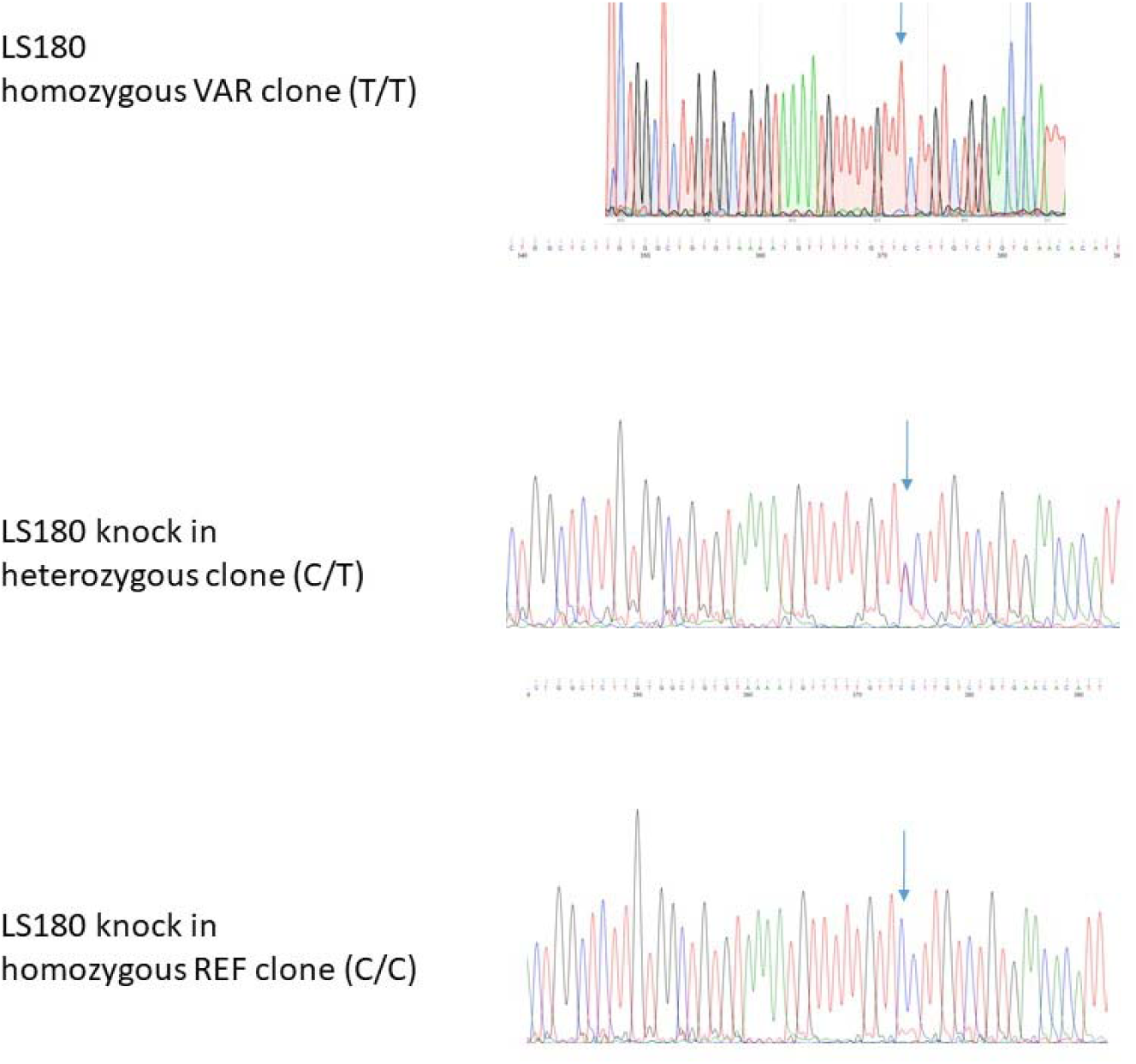
Sanger sequencing of *NXPE1* promoter site for LS180 knock in clones where SNP rs661946 (indicated by blue arrow, Hg38 chr11:114,559,887) has been converted to C/T (heterozygous) and C/C (homozygous REF). LS180 parent cell line is T/T (homozygous VAR) and is shown on top.

**Figure S9:**
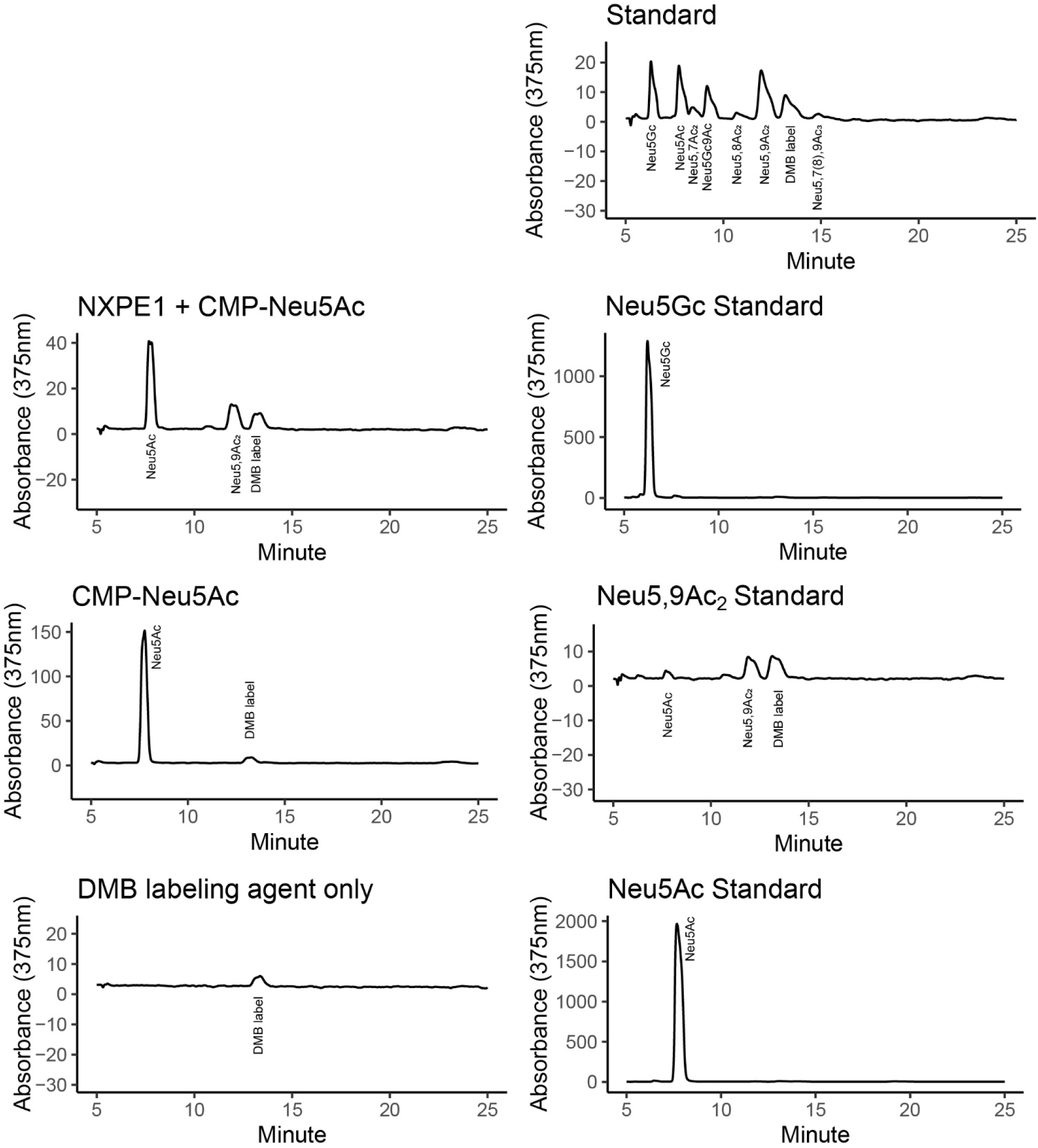
HPLC spectrum showing DMB-based HPLC readout of an enzymatic reaction containing CMP-Neu5Ac and acetyl coenzyme A, with or without the predicted extracellular domain of NXPE1 (AA 60-547). Note that some of the images above are also shown in figure 5A. N-glycolylneuraminic acid (Neu5Gc) is a sialic acid not found in humans and is shown as a control.

